# Clinical Characteristics of SARS-CoV-2 Pneumonia Compared to Controls in Chinese Han Population

**DOI:** 10.1101/2020.03.08.20031658

**Authors:** Yang Xu, Yi-rong Li, Qiang Zeng, Zhi-bing Lu, Yong-zhe Li, Wei Wu, Sheng-yong Dong, Gang Huang, Xing-huan Wang

## Abstract

**Background:** In December 2019, novel coronavirus (SARS-CoV-2) infected pneumonia occurred in Wuhan, China. The number of cases has increased rapidly but information on the clinical characteristics of SARS-CoV-2 pneumonia without comorbidities compared to normal controls in Chinese Han population is limited. Our objective is to describe the epidemiological and clinical characteristics of SARS-CoV-2 pneumonia without comorbidities compared to normal controls in the Chinese Han population.

**Methods:** Retrospective, multi-center case series of the 69 consecutive hospitalized patients with confirmed SARS-CoV-2 pneumonia, from February 7 to February 28, 2020; final date of follow-up was February 29, 2020.

**Results:** The study population included 69 hospitalized patients with confirmed SARS-CoV-2 pneumonia without comorbidities and 14,117 normal controls. 50.7% patients were male and 49.3% were female; 1.5% patients were asymptomatic cases, 63.8% patients were mild cases, and 36.2% patients were severe or critical cases. Compared with mild patients (n=44), severe or critical patients (n=25) were significantly older (median age, 67 years [IQR, 58-79] vs. 49 years [IQR, 36-60]; *p*<0.01). Fever was present in 98.6% of the patients. The second most common symptom was cough (62.3%), fatigue (58.0%), sputum (39.1%), and headache (33.3%). The median incubation period was 4 days (IQR, 2 to 7). Leukocyte count was 74.1% of normal controls and lymphocyte count was 45.9% of normal controls. The phenomenon of lymphocyte depletion (PLD) observed in severe or critical cases in 100%. Levels of lactate dehydrogenase, D-dimer, procalcitonin, and interleukin-6 were showed significant differences between mild and severe or critical cases. Chest computed tomographic scans showed bilateral patchy patterns (49.3%), local patchy shadowing (29.0%), and ground glass opacity (21.7%). 7.3% patients were diagnosed ARDS, 7.3% patients were diagnosed acute cardiac injury (troponin I >28 pg/mL) and 4.4% patients were diagnosed fungal infections or shock. 4.3% patients have been discharged; 1.5% patient had died; 1.5% patient had recovery.

**Conclusions:** In this multicenter case series of 69 patients without comorbidities, the full spectrum of asymptomatic, mild, severe, and critical cases is described. 50.7% patients were male and 49.3% were female; 1.5% patients were asymptomatic cases, 63.8% patients were mild cases, and 36.2% patients were severe or critical cases. 4.3% patients have been discharged; 1.5% patient had died; 1.5% patient had recovery. Among the 25 patients with severe or critical disease, 12.0% patients were underwent non-invasive mechanical ventilation, 8.0% patients underwent invasive mechanical ventilation, and 4.0% patients died.

## Introduction

In December 2019, a cluster of acute respiratory illness, now known as SARS-CoV-2 pneumonia, occurred in Wuhan, China.^1-5^ The disease has rapidly spread from Wuhan to other areas. As of February 11, 2020, the Chinese Center for Disease Control and Prevention (China CDC) has officially reported that there are 2.0% (889) asymptomatic cases, 2.3% (1,023) death cases, and 80.9% mild cases among 44,672 confirmed cases. 51.4% (22,981) were male and 48.6% (21,691) were female.^6^

In January, 2020, the 2019 novel coronavirus (SARS-CoV-2) was identified in samples of bronchoalveolar lavage fluid from a patient in Wuhan and was confirmed as the cause of the SARS-CoV-2 pneumonia.^1,2^ Full-genome sequencing and phylogenic analysis indicated that SARS-CoV-2 is a distinct clade from the betacoronaviruses associated with human severe acute respiratory syndrome (SARS) and Middle East respiratory syndrome (MERS). ^1,2^

COVID-19 has spread rapidly since it was first identified in Wuhan and has been shown to have a wide spectrum of severity. Recently, a report shows that SARS-CoV and SARS-CoV-2 shared the same functional host-cell receptor, angiotensin-converting enzyme 2 (ACE2).^7^ Furthermore, SARS-CoV-2 binds to ACE2 receptors in 10-20 fold higher affinity than SARS-CoV binds to the same receptors.^7^ Another report shows SARS-CoV-2 cell entry depends on ACE2 and TMPRSS2.^8^ Since ACE2 receptors on lung alveolar epithelial cells and enterocytes of the small intestine are dominant, lung alveolar epithelial cells or enterocytes of the small intestine may be an important susceptibility factor for human.^9^

According to World Health Organization interim guidance on January 12, 2020, SARS-CoV-2 infection is classified as asymptomatic case, mild and severe cases of pneumonia, and critical cases of pneumonia (ARDS, sepsis, septic shock). Severe cases of pneumonia are defined as patients with respiratory rate > 30 breaths/min, severe respiratory distress, or SpO2 < 90% on room air.^10^ Asymptomatic case has been reported in China and Germany.^11,12^ Huang et al^3^ first reported 41 cases of SARS-CoV-2 pneumonia in which most patients had a history of exposure to Huanan Seafood Wholesale Market. Organ dysfunction (shock, acute respiratory distress syndrome [ARDS], acute cardiac injury, and acute kidney injury, etc.) and death can occur in severe or critical cases. Guan et al^4^ reported findings from 1099 cases of SARS-CoV-2 pneumonia and the results suggested that the SARS-CoV-2 infection clustered within groups of humans in close contact, and was more likely to affect older men with comorbidities. However, the full spectrum of disease without comorbidities or related to cellular immune functions is not yet known. The objective of this case series was to describe the clinical characteristics of SARS-CoV-2 pneumonia without comorbidities compared to 14,117 normal controls in Chinese Han population.

## Methods

### Study Design and Participants

This multicentre, retrospective study was done at Zhongnan Hospital of Wuhan University, Chinese PLA General Hospital, Peking Union Medical College Hospital, and affiliated hospitals of Shanghai University of Medicine & Health Sciences. This case series was approved by the institutional ethics board of Zhongnan Hospital of Wuhan University (No. 2020020) and Peking Union Medical College Hospital (ZS-1830). We extracted 1532 case data from China CDC in Shanghai (338 cases), Beijing (414 cases), and Wuhan (780 cases) with clinical confirmed COVID-19. 1463 cases with comorbidities were excluded, including cardiovascular diseases (307 cases), hypertension (258 cases), chronic gastrointestinal diseases (126 cases),chronic renal diseases (125 cases), diabetes (122 cases), chronic obstructive pulmonary diseases (105 cases), hepatitis B carriers (95 cases), depression (74 cases), autoimmune diseases (73 cases), tumors (21 cases) and other diseases (157 cases). All 69 consecutive patients with confirmed COVID-19 without comorbidities from February 7 to February 28, 2020 and 14,117 normal controls without comorbidities from health checkup (from November 2018 to November 2019, before COVID-19 outbreak) in Chinese PLA General Hospital, Peking Union Medical College Hospital, Zhongnan Hospital of Wuhan University, and affiliated hospitals of Shanghai University of Medicine & Health Sciences were retrospectively enrolled. To reduce the impact of comorbidities on our research, all patients with comorbidities described above were excluded from the study. Identification of patients was achieved by reviewing and analyzing available electronic medical records and patient care resources. Written informed consent was waived due to the rapid emergence of this infectious disease. We retrospectively analyzed patients according to WHO interim guidance.^13^ Laboratory confirmation of SARS-CoV-2 infection was performed as previously described.^14^

All patients with SARS-CoV-2 pneumonia enrolled in this study were diagnosed according to World Health Organization interim guidance.^13^ According to World Health Organization interim guidance on January 12, 2020, SARS-CoV-2 infection is classified as asymptomatic cases, mild and severe cases of pneumonia, and critical cases of pneumonia (ARDS, sepsis, septic shock). Severe case of pneumonia is defined as patients with respiratory rate > 30 breaths/min, severe respiratory distress, or SpO2 < 90% on room air.^10^ Clinical outcomes (discharges, mortality, and recovery, etc.) were monitored up to February 29, 2020, the final date of follow-up. Cardiac injury was defined if the serum levels of cardiac biomarkers (eg, troponin I) were above the 99th percentile upper reference limit (>28 pg/mL) or new abnormalities were shown in electrocardiography and echocardiography.^3^

### Real-Time Reverse Transcription Polymerase Chain Reaction Assay for SARS-CoV-2

A confirmed case of COVID-19 is defined as a positive result on high throughput sequencing or real-time reverse-transcriptase–polymerase-chain-reaction (RT-PCR) assay of pharyngeal swab specimens. Samples were collected for extracting SARS-CoV-2 RNA from patients suspected of having SARS-CoV-2 infection as described previously.^14^ In brief, the pharyngeal swabs were placed into a collection tube with 150 μL of virus preservation solution, and total RNA was extracted within 2 hours using the respiratory sample RNA isolation kit. Forty μL of cell lysates were transferred into a collection tube followed by vortex for 10 seconds. After standing at room temperature for 10 minutes, the collection tube was centrifugated at 1000 rpm/min for 5 minutes. The suspension was used for RT-PCR assay of SARS-CoV-2 RNA. Two target genes, including open reading frame 1ab (*ORF1ab*) and nucleocapsid protein (N), were simultaneously amplified and tested during the real-time RT-PCR assay. Target 1 (*ORF1ab*): forward primer CCCTGTGGGTTTTACACTTAA; reverse primer ACGATTGTGCATCAGCTGA; and the probe 5′-VIC-CCGTCTGCGGTATGTGGAAAGGTTATGG-BHQ1-3′. Target 2 (N): forward primer GGGGAACTTCTCCTGCTAGAAT; reverse primer CAGACATTTTGCTCTCAAGCTG; and the probe 5′-FAM-TTGCTGCTGCTTGACAGATT-TAMRA-3′. The real-time RT-PCR assay was performed using a SARS-CoV-2 nucleic acid detection kit. Reaction mixture contains 12 μL of reaction buffer, 4 μL of enzyme solution, 4 μL of probe primers solution, 3 μL of diethyl pyrocarbonate–treated water, and 2 μL of RNA template. RT-PCR assay was performed under the following conditions: incubation at 50 °C for 15 minutes and 95 °C for 5 minutes, 40 cycles of denaturation at 94 °C for 15 seconds, and extending and collecting fluorescence signal at 55 °C for 45 seconds. A cycle threshold value (Ct-value) less than 37 was defined as a positive test result, and a Ct-value of 40 or more was defined as a negative test. These diagnostic criteria were based on the recommendation by the National Institute for Viral Disease Control and Prevention (China). A medium load, defined as a Ct-value of 37 to less than 40, required confirmation by retesting. Only RT-PCR confirmed cases were included in the analysis.

### Flow Cytometry Data Analysis

All antibodies were obtained from BD Biosciences (San Jose, CA, USA). Two 100-μL samples of blood were placed in two tubes for staining according to mmanufacturer’s manual. After this procedure, 1 mL of red-cell lyses buffer was added to each tube and incubated for 10 minutes and then washed with Sorvall cell washer (Thermo Fisher Scientific, Waltham, MA, USA). Cells were then resuspended in 350 μL of phosphate-buffered saline and acquired with the use of FACSCanto*(tm)* flow cytometry; daily quality control and assurance were carried out with the use of seven-color setup beads (BD Bioscience, San Jose, CA, USA).

### Statistical Analysis

Categorical variables were described as frequency rates and percentages, and continuous variables were described using mean, median, and interquartile range (IQR) values. Means for continuous variables were compared using independent group *t* tests when the data were normally distributed; otherwise, the Mann-Whitney test was used. Data (non-normal distribution) from repeated measures were compared using the generalized linear mixed model. Proportions for categorical variables were compared using the χ^2^ test, although the Fisher exact test was used when the data were limited. All statistical analyses were performed using SPSS (Statistical Package for the Social Sciences) version 13.0 software (SPSS Inc). For unadjusted comparisons, a 2-sided α of less than 0.05 was considered statistically significant. The analyses have not been adjusted for multiple comparisons and, given the potential for type I error, the findings should be interpreted as exploratory and descriptive. Because the cohort of patients in our study was not derived from random selection, all statistics are deemed to be descriptive only.

## Results

### Presenting Characteristics

The study population included 69 hospitalized patients with confirmed SARS-CoV-2 pneumonia without comorbidities and 14,117 normal controls. 50.7% patients were male and 49.3% were female; 1.5% patients were asymptomatic cases, 63.8% (44/69) patients were mild cases, and 36.2% (25/69) patients were severe or critical cases. Compared with mild patients (n=44), severe or critical patients (n=25) were significantly older (median age, 67 years [IQR, 58-79] vs. 49 years [IQR, 36-60]; *p*<0.01). Fever was present in 98.6% of the patients. The second most common symptom was cough (62.3%), fatigue (58.0%), sputum (39.1%), and headache (33.3%). Less common symptoms were chill, sore throat, shortness of breath, anorexia, diarrhea, nausea, and vomiting (Table 1). The median incubation period was 4 days (IQR, 2 to 7). On admission, the degree of severity of COVID-19 was categorized as mild in 44 patients and severe or critical in 25 patients. Compared with mild patients (n=44), severe or critical patients (n=25) were significantly older (median age, 67 years [IQR, 58-79] vs. 49 years [IQR, 36-60]; *p*<0.01) (Table 1).

**Table 1.** Demographics and Characteristics of 69 COVID-19 patients without comorbidities.

| Contents | All patients<br>(n=69) | Mild cases<br>(n=44) | Severe or critical<br>cases (n=25) | p value |
| --- | --- | --- | --- | --- |
| **Clinical characteristics** |  |  |  |  |
| Age, median (IQR), y | 57 (43-69) | 49 (36-60) | 67 (58-79) | 0.007 |
| < 59 | 42 (60.9%) | 32 (72.7%) | 10 (40%) |  |
| >60 | 27 (39.1%) | 12 (27.3%) | 15 (60%) |  |
| Sex |  |  |  |  |
| Male | 35 (50.7%) | 22 (50%) | 13 (52%) | 0.873 |
| Female | 34 (49.3%) | 22 (50%) | 12 (48%) |  |
| Current smoking | 5 (7.2%) | 2 (4.6%) | 3 (12.0%) | 0.344 |
| **Main symptoms** |  |  |  |  |
| Fever | 68 (98.6%) | 43 (97.7%) | 25 (100%) | 1.000 |
| Cough | 43 (62.3%) | 24 (54.6%) | 19 (76%) | 0.077 |
| Fatigue | 40 (58.0%) | 22 (50.0%) | 18 (72.0%) | 0.075 |
| Sputum | 27 (39.1%) | 16 (36.4%) | 11 (44.0%) | 0.532 |
| Chill | 10 (14.5%) | 4 (9.1%) | 6 (24.0%) | 0.152 |
| Sore throat | 10 (14.5%) | 5 (11.4%) | 5 (20.0%) | 0.478 |
| Shortness of breath | 11 (15.9%) | 2 (4.6%) | 9 (36.0%) | 0.001 |
| Headache | 23 (33.3%) | 18 (40.9%) | 5 (20.0%) | 0.077 |
| Anorexia | 8 (11.6%) | 4 (9.0%) | 4 (16%) | 0.448 |
| Diarrhea | 7 (10.1%) | 5 (11.4%) | 2 (8.0%) | 1.000 |
| Nausea or vomiting | 6 (8.7%) | 4 (9.0%) | 2 (8.0%) | 1.000 |
| Median incubation period | 4 (2-7) | 4 (2.5-7) | 4 (2-7) |  |
| **Laboratory characteristics** |  |  |  |  |
| White blood cell count, 10<sup>9</sup>/L | 4.3 (3.5-5.1) | 4.2 (3.2-5.0) | 4.6 (3.5-5.7) | 0.195 |
| Neutrophil count, 10<sup>9</sup>/L | 2.6 (2.0-3.6) | 2.3 (1.8-3.3) | 2.9 (2.4-4.6) | 0.021 |
| Lymphocyte count, 10<sup>9</sup>/L | 0.9 (0.7-1.2) | 1.2 (1.0-1.5) | 0.8 (0.6-1.0) | 0.031 |
| Monocyte count, 10<sup>9</sup>/L | 0.3 (0.2-0.4) | 0.3 (0.2-0.4) | 0.3 (0.2-0.4) | 0.701 |
| Hemoglobin, g/L | 138.6 (127.1-151.6) | 138.9 (126.4-151.6) | 138.2 (128.1-149.2) | 0.799 |
| Platelet count, 10<sup>9</sup>/L | 165.1 (135.3-218.7) | 168.1 (152.1-220.6) | 156.1 (121.3-217.7) | 0.357 |
| Prothrombin time, s | 12.1 (11.0-13.2) | 12.2 (11.0-13.2) | 12.3 (11.5-12.7) | 0.765 |
| Creatinine, µmol/L | 61.1 (47.3-69.7) | 57.1 (46.8-68.9) | 63.9 (51.1-79.3) | 0.179 |
| ALT*, U/L | 19.2 (15.1-33.1) | 18.2 (14.3-32.1) | 24.6 (15.1-35.2) | 0.297 |
| AST*, U/L | 33.8 (26.1-47.8) | 32.9 (23.9-41.1) | 45.2 (30.5-69.2) | 0.127 |
| Creatinine kinase, U/L | 94.1 (57.2-138.1) | 92.1 (57.2-134.2) | 118.7 (54.9-187.6) | 0.581 |
| Lactate dehydrogenase, U/L | 287.0 (242.0-407.0) | 281.0 (232.0-352.0) | 398 (321.0-571.0) | 0.005 |
| Hypersensitive troponin I, pg/mL | 3.3 (1.1-8.5) | 3.5 (0.7-5.6) | 3.7 (3.2-161.0) | 0.045 |
| D-dimer, mg/L | 0.5 (0.3-1.2) | 0.5 (0.3-0.9) | 2.3 (0.6-14.1) | 0.005 |
| Procalcitonin, ng/mL | 0.1 (0.1-0.1) | 0.1 (0.1-0.1) | 0.1 (0.1-0.3) | 0.032 |
| CRP, mg/L | 12.1 (3.6-28.1) | 10.1 (3.5-21.2) | 25.3 (2.9-72.1) | 0.187 |
| Interleukin-6, pg/mL | 7.9 (6.2-10.7) | 5.9 (2.8-10.9) | 14.8 (7.5-45.3) | 0.009 |
| **CT evidence of pneumonia** |  |  |  |  |
| Bilateral patchy patterns | 34 (49.3%) | 26 (59.1%) | 8 (32.0%) |  |
| Ground glass opacity | 15 (21.7%) | 7 (15.9%) | 8 (32.0%) |  |
| Local patchy shadowing | 20 (29.0%) | 11 (25.0%) | 9 (36.0%) |  |
| **Complications** |  |  |  |  |
| ARDS* | 5 (7.3%) | 1 (2.3%) | 4 (16%) | 0.054 |
| Acute cardiac injury | 5 (7.3%) | 1 (2.3%) | 4 (16%) | 0.054 |
| Fungal infections | 3 (4.4%) | 0 | 3 (12%) | 0.044 |
| Shock | 3 (4.4%) | 0 | 3 (12%) | 0.044 |
| **Critical management** |  |  |  |  |
| Nasal cannula | 35 (50.7%) | 11 (25.0%) | 24 (96.0%) | <0.001 |
| Noninvasive mechanical ventilation | 3 (4.4%) | 0 | 3 (12%) | 0.044 |
| Invasive mechanical ventilation | 2 (2.9%) | 0 | 2 (8.0%) | 0.128 |
| ECMO* | 0 | 0 | 0 |  |
| **Treatment** |  |  |  |  |
| Antiviral therapy | 38 (55.1%) | 22 (50.0%) | 16 (64.0%) | 0.261 |
| Antibiotic therapy | 31 (44.9%) | 11 (25.0%) | 20 (80%) | <0.001 |
| Antifungal therapy | 1 (1.5%) | 0 | 1 (4.0%) | 0.362 |
| Use of corticosteroid | 6 (8.7%) | 0 | 6 (24.0%) | 0.001 |
| **Prognosis** |  |  |  |  |
| Discharge | 3 (4.3%) | 3 (6.8%) | 0 (0%) | 0.024 |
| Death | 1 (1.5%) | 0 | 1 (4.0%) | 0.362 |
| Recovery | 1 (1.5%) | 1 (4.0%) | 0 | 0.362 |

\*ALT: alanine aminotransferase, AST: aspartate aminotransferase, CRP: C-reactive protein, ARDS: acute respiratory distress syndrome, ECMO: extracorporeal membrane oxygenation.

The COVID-19 mortality rate in China is 2.3%.^6^ However, the mortality rate is 4.6% (2,282 death cases in 49,540 confirmed cases)in Wuhan, 1.9% (8 death cases in 414 confirmed cases) in Beijing, and 0.9% (3 death cases in 338 confirmed cases) in Shanghai based on China CDC official report on February 29, 2020. Our findings showed that the levels of lymphocytes were 0.8(IQR, 0.6-1.1)10^9^/L in Wuhan, 1.0(IQR, 0.7-1.4)10^9^/L in Beijing, and 1.1 (IQR, 0.8-1.5) 10^9^/L in Shanghai before admission to hospitals, respectively, indicating that cellular immune function related to the severity of disease. Our previous report shows that neutrophils increase in non-survivors on day 11 and day 17, whereas, lymphocytes decrease in non-survivors on day 11 and day 17 significantly (*p* < 0.05) indicating cellular immunity related to mortality.^14^

In 69 patients, there was an asymptomatic case in admission and the patient developed symptom 3 days after positive RT-PCR test.

### Radiologic and Laboratory Parameters in Mild and Severe or Critical Patients

The radiologic and laboratory findings were showed (Table 1). These data could helpfully distinguish between mild and severe or critical patients. All of the 69 enrolled patients showed bilateral patchy patterns, or ground-glass opacity, or local patchy shadowing of chest CT scan. (Table 1, Figure 1). On admission, the predominant pattern of abnormality observed was bilateral patchy patterns (49.3%), local patchy shadowing (29.0%), and ground glass opacity (21.7%).

**Figure 1.**
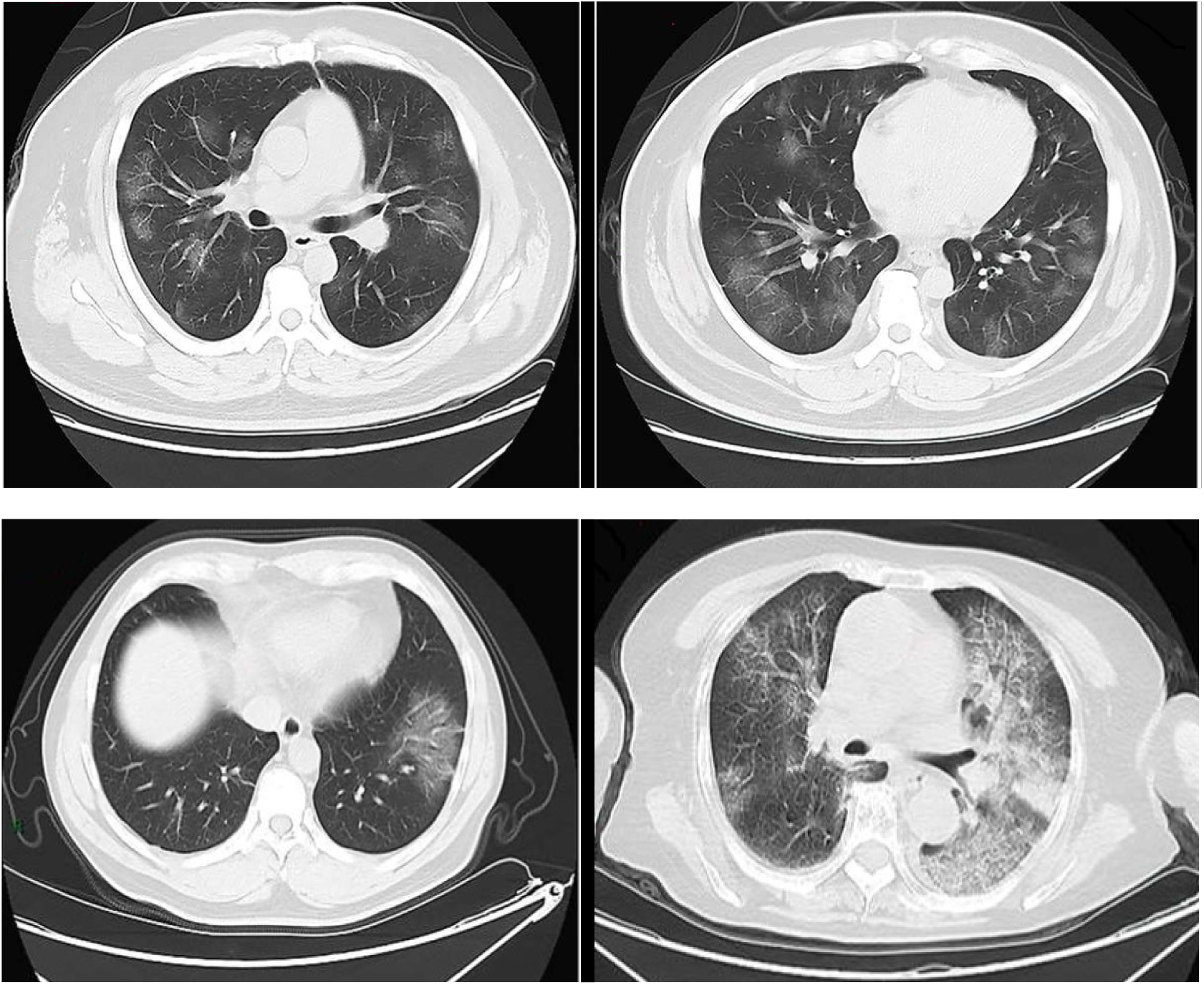
Representative chest radiographic manifestations in mild and severe cases with COVID-19. Chest computed tomography imaging from mild cases (top two imaging) and from severe cases (bottom two imaging).

Due to lack of reference ranges for blood routine and lymphocytes of normal Chinese Han population, we systematically analyzed the 14,117 healthy adult Chinese Han people of 18-86 years old and classified them as an age group every 10 years for the convenience of other researchers in the future (Table 2).

**Table 2.** Laboratory characteristics of 14,117 normal controls.

| **Contents** | **Units** | **<20**<br>**(n = 31)** | **20-29**<br>**(n = 444)** | **30-39**<br>**(n = 1766)** | **40-49**<br>**(n = 5781)** |
| --- | --- | --- | --- | --- | --- |
| White blood cell | 10<sup>9</sup>/L | 6.50<br>(5.35-8.07) | 6.26<br>(5.16-7.29) | 5.98<br>(5.06-7.06) | 5.78<br>(4.94-6.82) |
| Lymphocyte | cells/m<br>m<sup>3</sup> | 2444<br>(2276-2656) | 2223<br>(1860-2580) | 2041<br>(1679-2490) | 1941<br>(1608-2339) |
| Hemoglobin | g/L | 154<br>(142-157) | 140<br>(130-156) | 145<br>(130-156) | 144<br>(130-155) |
| Platelet | 10<sup>9</sup>/L | 236<br>(202-296) | 242<br>(206-282) | 240<br>(205-278) | 232<br>(198- 269) |
| Alanine<br>aminotransferase | U/L | 17<br>(11-29) | 15<br>(10-28) | 18<br>(12-30) | 18<br>(13-27) |
| Aspartate<br>aminotransferase | U/L | 17<br>(15-22) | 17<br>(14-21) | 17<br>(15-22) | 18<br>(15-21) |
| Creatinine | μmol/L | 66<br>(60-79) | 63<br>(54- 75) | 65<br>(54-75) | 66<br>(56-76) |
| C-reactive protein | mg/L | 0.12<br>(0.07-0.20) | 0.10<br>(0.05-0.18) | 0.10<br>(0.06-0.16) | 0.09<br>(0.05-0.16) |
| Lactate<br>dehydrogenase | U/L | 151<br>(138-162) | 143<br>(127-160) | 142<br>(127-159) | 144<br>(130-160) |

| **Contents** | **Units** | **50-59**<br>**(n = 4801)** | **60-69**<br>**(n = 1046)** | **70-79**<br>**(n = 227)** | **>80**<br>**(n = 21)** |
| --- | --- | --- | --- | --- | --- |
| White blood cell | 10<sup>9</sup>/L | 5.54<br>(4.72-6.46) | 5.52<br>(4.69-6.42) | 5.70<br>(4.73-6.40) | 4.83<br>(4.45-6.19) |
| Lymphocyte | cells/m<br>m<sup>3</sup> | 1920<br>(1584-2289) | 1841<br>(1529-2239) | 1781<br>(1404-2241) | 1456<br>(1242-1790) |
| Hemoglobin | g/L | 143<br>(131-154) | 140<br>(131-151) | 137<br>(129-146) | 140<br>(127-154) |
| Platelet | 10<sup>9</sup>/L | 223<br>(190-260) | 213<br>(179-248) | 210<br>(174-250) | 189<br>(176-224) |
| Alanine<br>aminotransferase | U/L | 18<br>(14-25) | 16<br>(13-23) | 14<br>(11-18) | 14<br>(10-22) |
| Aspartate<br>aminotransferase | U/L | 18<br>(15-22) | 18<br>(16-22) | 18<br>(15-21) | 21<br>(17-29) |
| Creatinine | μmol/L | 66<br>(56-76) | 64<br>(56-75) | 65<br>(56-74) | 70<br>(64-75) |
| C-reactive protein | mg/L | 0.09<br>(0.05-0.16) | 0.10<br>(0.06-0.18) | 0.12<br>(0.07-0.22) | 0.10<br>(0.08-0.17) |
| Lactate<br>dehydrogenase | U/L | 151<br>(135-170) | 158<br>(140-177) | 160<br>(144-181) | 165<br>(148-200) |

The reference ranges of blood routine are as followings (Table 2): white blood cell (4.83-6.50, 10^9^/L), lymphocyte (1.5-2.4, 10^9^/L), hemoglobin (140-154, g/L), platelet (189-236, 10^9^/L), alanine aminotransferase (14-18, U/L), aspartate aminotransferase (17-21, U/L), creatinine (63-70, μmol/L), C-reactive protein (0.09-0.12, mg/L), and lactate dehydrogenase (142-165, U/L).

On admission, leucocytes were no difference between mild and severe or critical cases (Table 1). However, the patient’s leukocyte count was 74.1% (4.3/5.8) of the median value in normal controls (Table 1, Table 2). Neutrophils and lymphocytes showed significant difference (*p* < 0.05) between mild and severe or critical cases (Table 1). In addition, the patient’s lymphocyte count was 45.9% (0.9/1.959) of the median value in normal controls (Table 1, Table 2).

The phenomenon of lymphocyte depletion (PLD) observed in severe or critical cases (Table 3). As the disease progressed and clinical status deteriorated, the levels of lymphocytes progressively decreased before death. Further analyzing 69 cases of lymphocyte subsets showed that CD4 and CD8 T lymphocytes have significant difference (*p* < 0.01) between mild and severe or critical cases (Table 4).

**Table 3.**
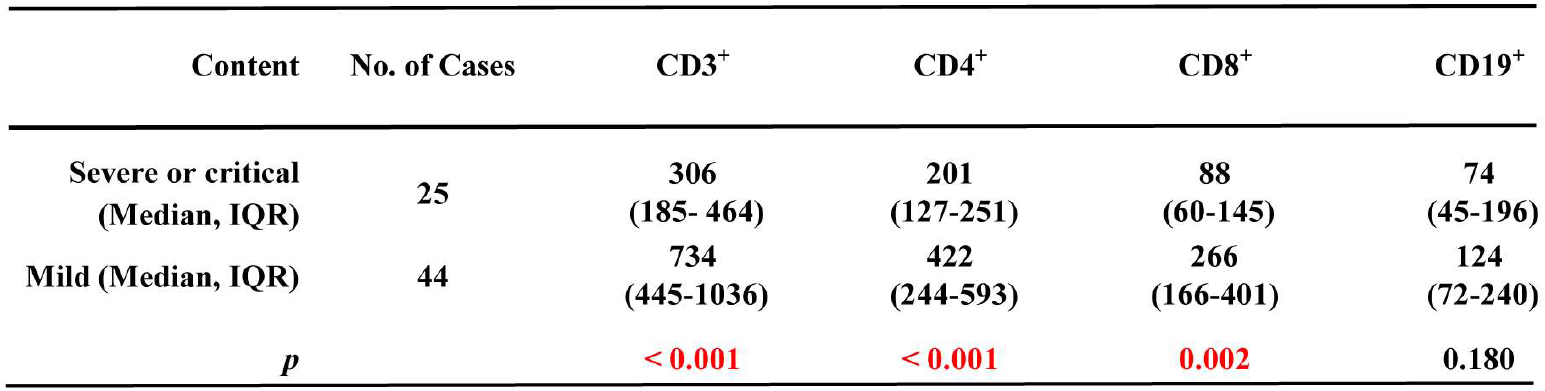
Lymphocyte subsets in COVID-19 pneumonia mild vs. severe or critical.

Our findings also show significant differences (*p* < 0.05) in lactate dehydrogenase, troponin I, D-dimer, procalcitonin, and interleukin-6 (IL6) between mild and severe or critical cases (Table 1). Levels of aspartate aminotransferase of patients were elevated in comparison with normal controls. The levels of C reaction protein (CRP) were 121-fold higher than the value in normal controls, however, no difference observed between mild and severe or critical cases (Table 1, Table 2). There are 3.6% patients with acute kidney injury, however, no difference observed between mild and severe cases.^14^ There is no acute kidney injury since serum creatinine level is within a normal range (Table 1, Table 2), indicating that patients with comorbidities might prone to acute kidney injury.^14^

All patients were tested for nine respiratory pathogens, the nucleic acid of influenza viruses A and B. We did not find other respiratory viruses in any of the patients. Three cases of fungal infections were diagnosed as *Candida albicans*.

### Main Interventions and Outcomes

All patients were treated in isolation. 38 (55.1%) patients received antiviral treatment, including oseltamivir (75 mg every 12 h, orally). 31 (44.9%) patients were treated with antibiotics. The antibiotics used generally covered mild pathogens and some atypical pathogens; when secondary bacterial infection occurred, medication was administered according to the results of bacterial culture and drug sensitivity. The antibiotics used were moxifloxacin, ceftriaxone, azithromycin, and tigecycline or linezolid against methicillin-resistant *Staphylococcus aureus*, as well as antifungal drugs. Six patients significantly increased in IL6 were also treated with methylprednisolone. Three patients used non-invasive ventilator mechanical ventilation. Two patients used an invasive ventilator to assist ventilation. The ventilator adopted P-SIMV mode, the inhaled oxygen concentration was 35–100%, and the positive end-expiratory pressure was 6–12 cm H2O. No patients were treated with extracorporeal membrane oxygenation (ECMO). Moreover, 7.3% patients were diagnosed ARDS, 7.3% patients were diagnosed acute cardiac injury (troponin I>28 pg/mL) and 4.4% patients were diagnosed fungal infections or shock (Table 1).

By the end of February 29, three (4.3%) patients have been discharged; one (1.5%) patient had died; one (1.5%) patient had recovery; 64 patients were still in hospital (Table 1). Among the 25 patients with severe or critical disease, a primary composite end-point event occurred in 5 patients (20.0%), including 12.0% who were underwent non-invasive mechanical ventilation, 8.0% who underwent invasive mechanical ventilation, and 4.0% who died (Table 1).

## Discussion

This report, to our knowledge, is the first retrospective multicenter study of COVID-19 with normal controls. The study population included 69 hospitalized COVID-19 patients without comorbidities and 14,117 normal controls. 50.7% patients were male and 49.3% were female; 1.5% patients were asymptomatic cases, 63.8% patients were mild cases, and 36.2% patients were severe or critical cases. The median incubation period was 4 days (IQR, 2 to 7). The median age of the patients was 57 years (IRQ, 43 to 69); 39.1% of the patients were older than 60 years of age. Fever was present in 98.6% of the patients. The second most common symptom was cough (62.3%), fatigue (58.0%), sputum (39.1%), and headache (33.3%). Less common symptoms were chill, sore throat, shortness of breath, anorexia, diarrhea, nausea, and vomiting (Table 1). On admission, the degree of severity of COVID-19 was categorized as mild in 44 patients and severe or critical in 25 patients. Compared with mild patients (n=44), severe or critical patients (n=25) were significantly older (median age, 67 years [IQR, 58-79] vs. 49 years [IQR, 36-60]; *p*<0.01). There is no acute kidney injury since serum creatinine level is within a normal range (Table 1, Table 2), indicating that patients with comorbidities might prone to acute kidney injury.^14^

For the asymptomatic case, we speculate that the patient is currently taking HIV medications and the medication is interfering with the results of RT-PCR. Further research and observation are needed.

Our findings included the PLD at presentation were associated with the severity of disease. The severe PLD or the acquired immunodeficiency (CD4 lymphocytes are below 200 cells/mm^3^) is common in HIV patients. When CD4 lymphocytes are below 200 cells/mm^3^, clinicians might pay attention to fungal infections. There were three cases of fungal infections seen in our study indicating the lymphocyte depletion associated with fungal infections (Table 1, Table 3) as patients with T cell depletion have increased susceptibility to fungal infections.^15,16^ Our findings also show significant differences (*p* < 0.01) in IL6 and D-dimer between mild and severe or critical groups, suggesting that IL6 and D-dimer are important parameters for the severity of disease (Table 1).

It has been reported that SARS-CoV directly infects monocytes, macrophages and T lymphocytes in human.^17^ The PLD may also be secondary to activation of T lymphocytes.^18^ COVID-19 has spread rapidly since it was first identified in Wuhan and has been shown to have a wide spectrum of severity as SARS-CoV-2 binds to ACE2 receptors in 10-20 fold higher affinity than SARS-CoV binds to the same receptors.^7,19^ The report shows SARS-CoV-2 cell entry depends on ACE2 and TMPRSS2.^8^ Therefore, it is necessary to further study whether lymphocytes have ACE2 receptor and TMPRSS2 expression during the developmental stage since the PLD exists. A SARS-CoV-2 RT-PCR assay for lymphocyte subsets in severe or critical cases should be followed up. Our findings also suggest that lymphocyte subsets should be analyzed at admission immediately no matter how COVID-19 affected lymphocytes through directly or indirectly. When patients are confirmed as COVID-19, clinicians should order tests for lymphocyte subsets in order to intervene early in the consequences of the PLD.

This study has several limitations. First, only 69 cases were included. However, this report is the first multicenter study of SARS-CoV-2 infection without comorbidities and 14,117 normal controls were analyzed. Through this report, a larger multicenter cohort study can be introduced in the future. Second, among the 69 cases, most patients are still hospitalized at the time of manuscript submission. Therefore, it is difficult to assess all risk factors for poor outcome. Third, the assay of IgG and IgM antibodies to SARS-CoV-2 in serum is not available yet. Fourth, the clinical characteristics of SARS-CoV-2 pneumonia of 2019 in Wuhan and SARS-CoV pneumonia of 2003 in Beijing should be compared and studied.^20^ Fifth, the pathophysiology of SARS-CoV-2 infecting the lungs and intestines through the ACE2 receptor and TMPRSS2 or the acquired immunodeficiency (CD4 cells below 200 cells/mm^3^) associated with COVID-19 (the severe or critical cases) seen in some patients directly or indirectly caused by SARS-CoV-2 is not understood, and further research and observation are needed.

## Conclusions

In this multicenter case series of 69 patients without comorbidities, the full spectrum of asymptomatic, mild, severe, and critical cases is described. 50.7% patients were male and 49.3% were female; 1.5% patients were asymptomatic cases, 63.8% patients were mild cases, and 36.2% patients were severe or critical cases. 4.3% patients have been discharged; 1.5% patient had died; 1.5% patient had recovery. Among the 25 patients with severe or critical disease, 12.0% patients were underwent non-invasive mechanical ventilation, 8.0% patients underwent invasive mechanical ventilation, and 4.0% patients died.

## Data Availability

we have the availability of all data.

